# Assessing Resistome Host Range Across Water Reclamation in Three Geographically Distinct Communities using Hi-C Sequencing

**DOI:** 10.64898/2026.02.12.26346186

**Authors:** Sarah E. Philo, Michael A. Saldana, Harmita Golwala, Siyi Zhou, Jeseth Delgado Vela, Lauren B. Stadler, Adam L. Smith

**Affiliations:** Sonny Astani Department of Civil and Environmental Engineering, University of Southern California, Los Angeles, California, USA; Department of Civil and Environmental Engineering, Rice University, Houston, Texas, USA; Department of Civil and Environmental Engineering, Duke University, Durham, North Carolina, USA

**Keywords:** antimicrobial resistance, proximity ligation, Hi-C, shotgun sequencing, wastewater surveillance

## Abstract

Antimicrobial resistance (AMR) is a growing problem, with annual deaths set to pass 10 million by 2050 if current trends continue. Wastewater surveillance has been proposed as a strategy to understand population-level resistance, and water reclamation facilities (WRFs) have been identified as a control point for environmental dissemination of resistant bacteria. Understanding dynamics of AMR across WRFs requires advanced molecular tools that elucidate host bacteria, especially for mobile resistance carried on plasmids. To that end, influent, activated sludge, and effluent were collected from three WRFs in North Carolina, Texas, and California during three weeks of Spring 2024. Samples were analyzed using Hi-C proximity ligation sequencing to identify the AMR host range for chromosomal and plasmid-based resistance. A total of 1,868 hits for 244 unique resistance genes were observed, with seven resistance genes identified in all samples. Resistance genes were more likely to be carried on a microbial plasmid in influent, but more likely to be in a chromosome in activated sludge. Seventeen total microbial hosts for resistance genes were identified in effluent, suggesting WRF effluents may be sources of resistant bacteria to receiving surface waters. A high proportion of all identified host relationships were confined to just four bacterial families. Hi-C contact mapping is a critical tool to more fully describe the AMR host range in complex matrices, particularly for plasmid-based resistance genes.

**Importance:** Antimicrobial resistance (AMR) threatens modern medicine. Water reclamation facilities receive a complex mixture of antibiotics and rely on active microbial communities for treatment, thereby acting as critical systems to prevent environmental spread of resistance. However, AMR dynamics are difficult to discern in complex wastewater environments due to antibiotic resistance genes (ARGs) being frequently carried on mobile pieces of DNA that are difficult to link to specific bacteria using conventional shotgun sequencing. Novel proximity ligation sample preparation techniques like Hi-C physically link co-located sequences of DNA before shotgun sequencing. This allows sequencing to elucidate the bacterial hosts for both stable and mobile ARGs. In the current study, Hi-C sequencing was carried out on influent, activated sludge, and effluent collected from water reclamation facilities in California, Texas, and North Carolina to assess the resistome host range across treatment.

**Graphical Abstract:** 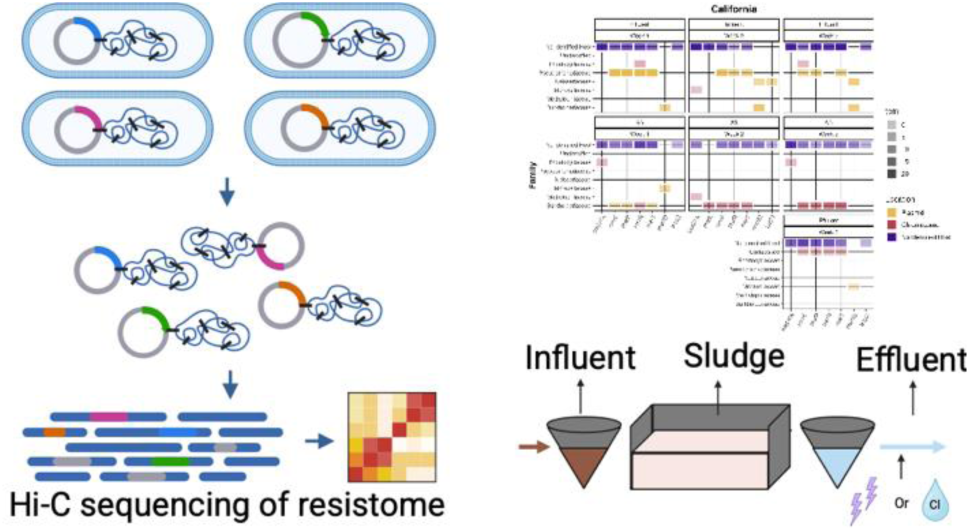

## 1 Introduction

Antimicrobial resistance (AMR) is one of the greatest threats to public health, with AMR associated deaths predicted to increase to 10 million deaths annually by 2050 from 4.9 million deaths in 2021 if growing resistance is not addressed (1). Despite increased attention to AMR, new resistance patterns are emerging worldwide. Most notably, plasmid-mediated colistin resistance (*mcr-1*) emerged in the early 2010s in China (2), and carbapenem-resistant Enterobacteriaceae (*bla*_NDM-1_) emerged in the early 2000s in India and Pakistan (3). Influent wastewater collected from water reclamation facilities (WRFs) has been proposed as a suitable sample to monitor population-level AMR because it is representative of the catchment population (4–6). A recent global study of influent wastewater identified 13 antibiotic resistance genes (ARGs) in every sample collected from 243 cities in 101 countries (i.e. the core resistome (7)), in addition to 127 ARGs in at least half of the samples (8). This suggests that wastewater can be used to identify similarities and differences in the resistome across geography. AMR surveillance across WRFs is additionally critical as these systems are considered resistance “hotspots” due to the combination of microbial communities, antibiotics, and disinfectants (9–11). ARGs are routinely detected in WRF effluents (12), making WRFs both sources of and critical control points to mitigate dissemination of resistance to the environment (13). Further, surface waters downstream of WRFs have been found to have higher ARG abundance relative to upstream surface waters (14–16), indicating the facilities themselves are sources of resistance in the environment.

There are numerous open questions about how different treatment processes in WRFs select for or remove AMR. A review by C. X. Hiller et al. (17), summarized the efficacy of different wastewater treatment processes, noting that antibiotic resistant bacteria (ARB) and ARGs often have different removal rates from the same treatment process. Critically, existing research does not identify if and how the bacteria carrying ARGs, i.e. the ARG host range, changes during treatment, particularly for non-culturable ARB. WRFs are designed to meet specific pathogen reduction values without considering ARB in effluents. Identifying drivers of the AMR host range throughout treatment and in effluents could decrease environmental risks of releasing ARBs.

Horizontal gene transfer (HGT) of resistance genes via mobile genetic elements (MGEs) and plasmids is particularly concerning because bacteria are often exposed to sub-lethal concentrations of antibiotics in WRFs, which has been shown to increase HGT rates (18). Understanding microbial hosts of resistance genes on plasmids is therefore crucial. Yet, shotgun metagenomic sequencing for wastewater-based AMR surveillance can only identify chromosomal microbial hosts of ARGs (4, 13, 19). The inability to link plasmid ARGs with host microbes remains a substantial weakness of shotgun sequencing to study AMR dynamics. Single-cell sequencing techniques like epicPCR have been developed to overcome these limitations (20). However, this method is a targeted approach limited to pre-chosen genes, preventing holistic analyses. Novel chromosome conformation capture (3C) techniques, such as Hi-C, link ARGs on plasmids to their host microbes by crosslinking physically close DNA in a non-targeted manner prior to cell lysis (21). These 3C techniques remain the only non-targeted methods to identify microbial-plasmid-ARG relationships, removing a substantial limitation of traditional shotgun sequencing for AMR surveillance. Hi-C has been employed to study AMR in untreated wastewater (21, 22) and across wastewater treatment to reconstruct plasmid and ARG networks in these matrices (23).

Numerous questions remain about AMR dynamics through different treatment trains, across geographic distance, and following disinfection. We hypothesize that 1) the relative proportion of ARGs with hosts will decrease across treatment, 2) the number of bacterial hosts will be different in the effluents from the three sampling locations due to different final treatment processes, and 3) a few bacterial families will be responsible for the bulk of the host relationships. In this study, we used Hi-C proximity-ligation sequencing to test these hypotheses by collecting influent, activated sludge (AS), and effluent from WRFs in California (CA), Texas (TX), and North Carolina (NC). We assessed AMR, the ARG host range, and how the resistome changed during wastewater treatment. Additionally, we assayed effluent to assess the risk of releasing clinically relevant ARB from WRFs.

## 2 Results

### 2.1 Bacterial community structure

Relative abundance of bacterial metagenome assembled genomes (MAGs, >70% completion, <20% contamination) was assessed at the class and family taxonomic levels (Figure 1). Results are only reported for six out of nine effluent samples collected as there was insufficient sequencing depth to complete the full analysis pipeline due to low DNA yields. MAGs were used to understand community structure because there is higher confidence in the sequencing quality and taxonomic assignment relative to the assemblies. The influent microbial communities in CA were highly stable week to week, while communities in NC and TX influent were less stable (Figure 1). The influent microbial communities in all three were dominated by *Gammaproteobacteria* (mean relative abundance 52%, 95% CI: 41 - 64%). However, microbial communities in AS from all three locations were highly stable week to week, exhibited strong similarity across geography, and were dominated by *Bacteroidia* spp. (mean relative abundance 42%, 95% CI: 28 – 56%, Figure 1). Effluent communities exhibited little similarity to each other over time or to the AS in all three sampling locations (Figure 1).

**Figure 1:**
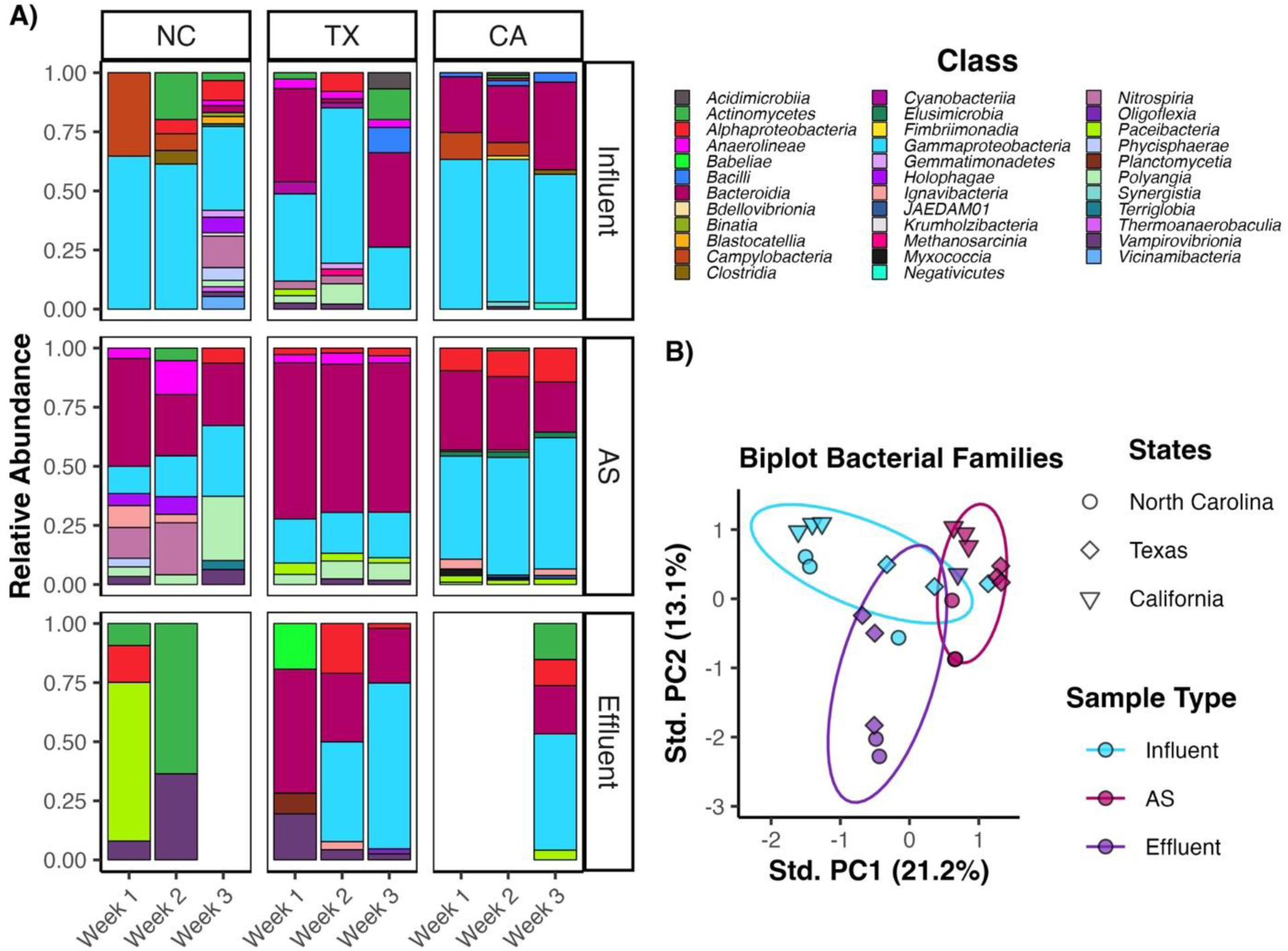
A) Relative abundance of bacterial classes for each sampling location, sample type, and sampling event of the MAGs. Effluent samples in NC and CA are not included because they could not undergo the full sequencing pipeline due to low mapping of Hi-C data to shotgun assemblies. Relative abundance was calculated with TPM adjusted to the microbial fraction of the shotgun sequencing data. B) Biplot of PCA with centering and scaling for bacterial family relative abundance following a Hellinger transformation of MAGs. MAGs were filtered for >70% completion and <20% contamination. Ellipses represent 68% coverage of the data for each sample type collected across geographic locations.

### 2.2 Resistome assessment

AMR was assessed at different levels (SI Figure 1). Here, total resistance genes was defined as the total count of resistance genes for drugs, heavy metals, and environmental stressors identified in AMRFinderPlus. There were 1,868 total resistance genes detected in the shotgun sequencing assemblies (Table 1, SI Figure 1). Total ARGs was defined as the subset of total resistance genes conferring resistance to solely pharmaceuticals (n = 955, Table 1). Unique total resistance genes and ARGs consisted of each individual gene detected and counted once. There were 244 unique total resistance genes and 173 unique ARGs (SI Figure 1). The number of total resistance genes detected was highest in the influent (*n* = 1099) compared to AS (*n* = 414). ARGs for pharmaceuticals similarly saw a reduction in detection from influent (n = 601) to AS (n = 192). Across all samples and gene types, the bulk of reduction occurred between the influent and AS (Table 1). Total resistance genes increased between AS and effluent in each week sampled for which there was an effluent sample (Table 1).

**Table 1:**
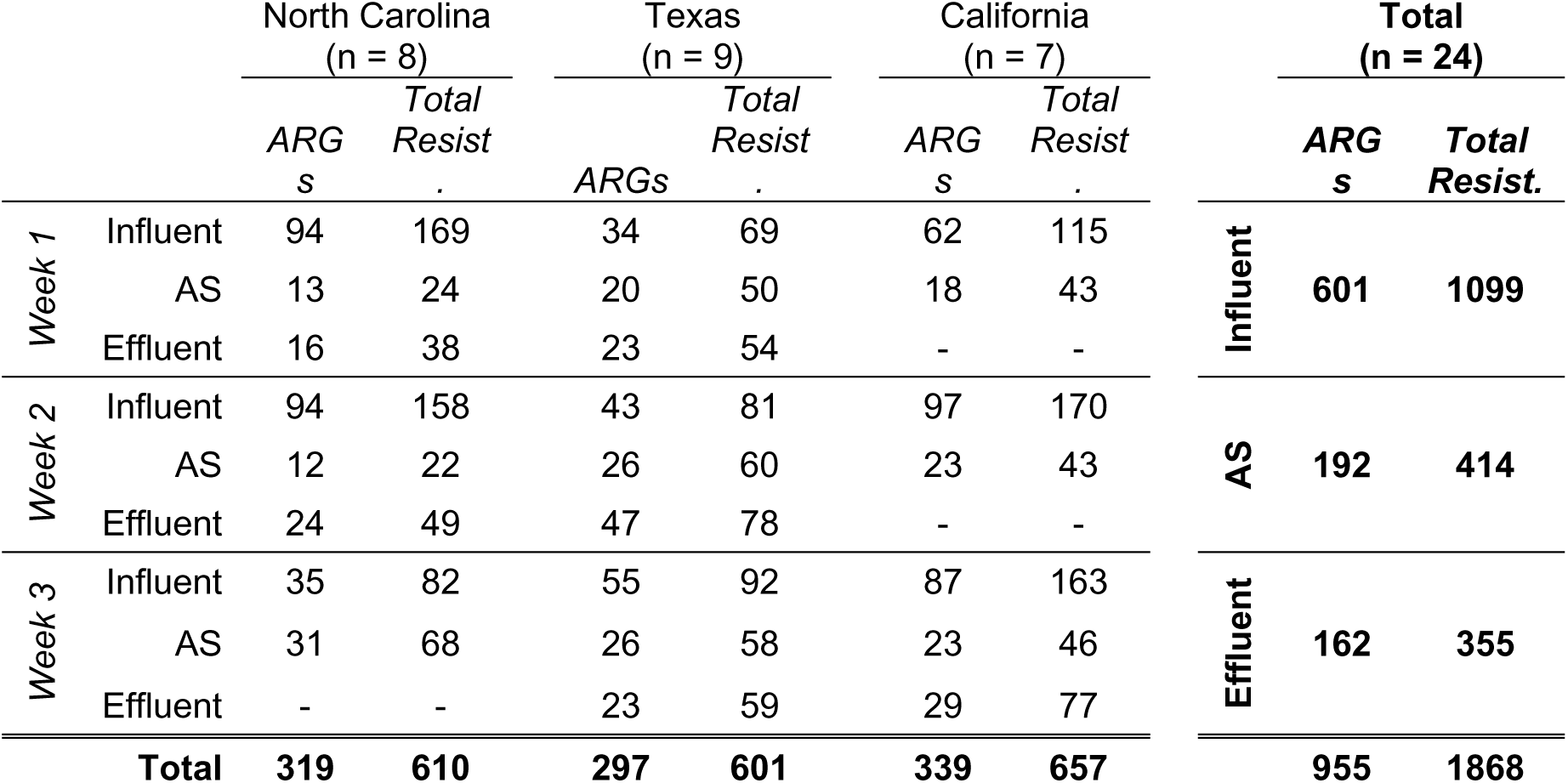
Total number of resistance genes detected in each sampling location at each treatment level. ARGs are representative only of resistance genes that confer resistance to pharmaceuticals. Total resistance includes both drug and heavy metal/environmental stressor resistance genes. The total number of samples analysed at each location is the n value.

The entire resistome including resistance genes for pharmaceuticals, heavy metals, and environmental stressors was assessed to understand how host carriage changes over treatment. When considering potential host bacteria, Hi-C sequencing identified plasmid-based hosts, chromosomal-based hosts, or no host. Hosts were assigned based on identification in a microbial genomic bin. In the event that no host was assigned, it was due to either 1) no informative Hi-C crosslinks between microbial bins and contigs containing resistance genes or 2) unbinned contigs leading to a lack of host identification for resistance genes (mean bin coverage = 42.4%, SI Table 1), which is a shared challenge with traditional shotgun sequencing analysis. While the majority of detected resistance genes did not have a microbial host identified across all sample types and locations, resistance genes were more likely to be in a microbial plasmid in influent and in a chromosome in AS (Table 2). Very few microbial hosts were identified in effluent samples (Table 2), despite increased detection of total resistance genes from AS to effluent in weeks where both samples underwent the full sequencing pipeline (Table 1).

**Table 2:**
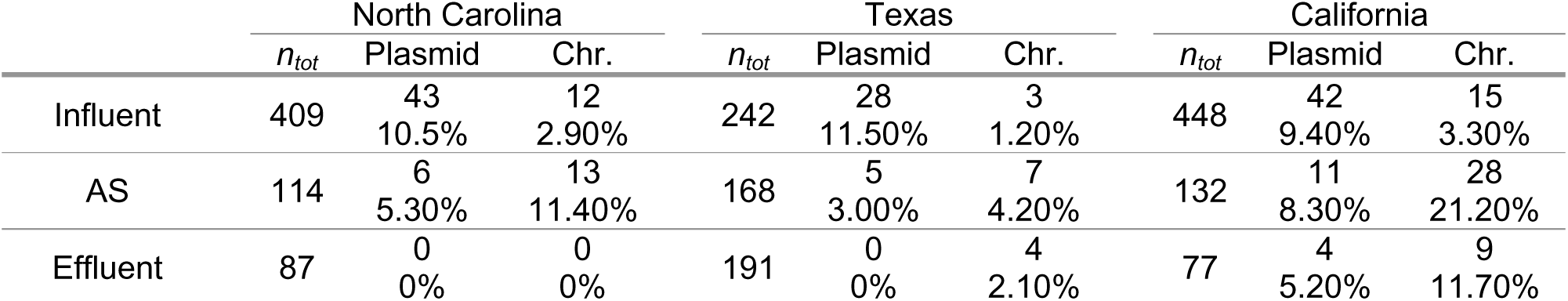
Total resistance genes detected in either a bacterial host plasmid or chromosome (Chr.) by Hi-C contact mapping across all samples that underwent the complete deconvolution platform. n_tot_ represents the total number of genes detected in that sample type from that geographic area. Percentages represent the fraction of genes detected in either a microbial plasmid or chromosome from that sample type and geographic location.

Among the ARGs, the most commonly detected antibiotic class across all samples collected was beta-lactams (*n* = 320), followed by macrolides (*n =* 180), tetracycline (*n* = 138), and aminoglycoside (*n* = 112) (SI Table 2). When considering relative abundance, macrolide resistance and beta-lactam resistance were the most abundant on average across all samples, with not statistically different relative abundances of 27.7% (95% CI = 25.1% - 30.4%) and 23.7% (95% CI = 21.5% - 26.0%), respectively (Figure 2A, SI Table 2). To assess resistome similarity across sampling locations, PCA was carried out on the relative abundance of antibiotic classes (Figure 2B, SI Figure 2). Although there was slight clustering by sample type (SI Figure 2), PCA did not identify any unique resistome clusters, as all sample ellipses overlapped with each other (Figure 2B), Resistance to specific antibiotic classes stayed relatively stable over time, between sampling locations, and across treatment processes (Figure 2). Additionally, even though different secondary treatment processes are employed at NC (5-stage Bardenpho) and TX and CA (high-purity O_2_ activated sludge), the resistance gene abundance in the AS from all three WRFs was highly similar (SI Figure 2).

**Figure 2:**
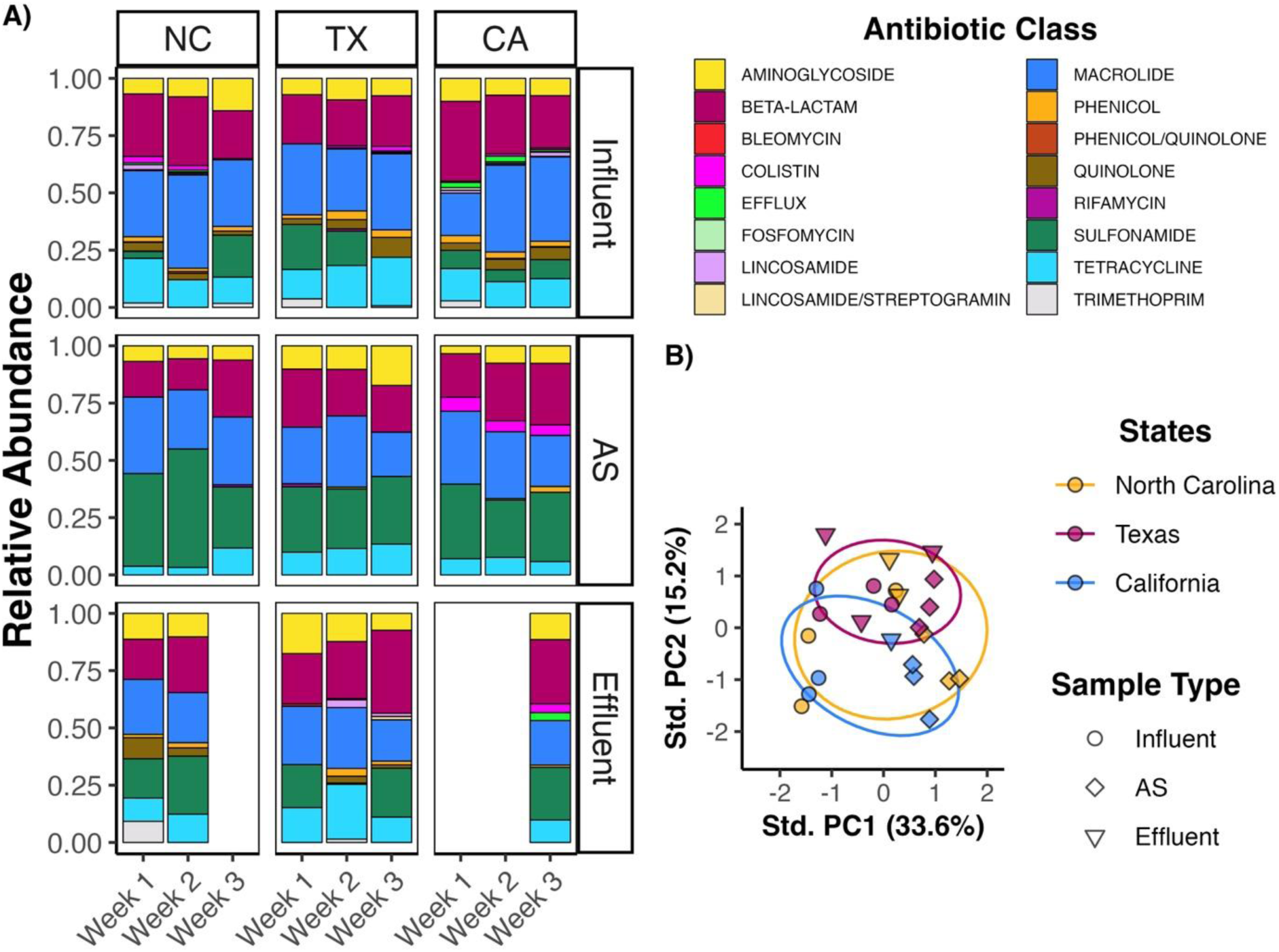
A) Relative abundance of antibiotic classes over treatment and time. Relative abundance represents the adjusted TPM for the ARG fraction of the shotgun sequencing data. B) Principal component analysis with centering and scaling of the resistome relative abundance at the antibiotic class level. Ellipses represent 68% coverage of the data for samples collected from each state. Prior to PCA, data underwent a Hellinger transformation. Effluent samples in NC and CA are not included because they could not undergo the full sequencing pipeline due to low mapping of Hi-C data to assemblies. Contigs were filtered for ARG sequences that confer resistance to pharmaceuticals.

### 2.3 Core Resistome

Seven resistance genes were identified in the core resistome: *bla*_OXA_*, merE, merP, merR, merT, msr(E), and tet(C)* (SI Table 3, Figure 3). They were detected a total of 505 times with 102 host relationships. Four genes, *merE, merP, merR,* and *merT,* encode for mercury resistance in the *mer* operon (24, 25), working together via disparate functions to detoxify mercury. The remaining three genes in the core resistome, *bla*_OXA_*, msr(E),* and *tet(C)* encode for resistance to drugs. *bla*_OXA_ encompasses a diverse group of genes encoding for class D β-lactamases and confer resistance against later generation cephalosporins and carbapenem (26). Additional *bla_OXA_*alleles detected were *bla_OXA-_*_1, 2, 4, 5, 9, 10, 119, 347, 732 and 912_, although no hosts were identified (SI Data). Hi-C crosslinking identified *bla_OXA_* as being carried on bacterial chromosomes in *Aeromonas caviae* in NC and TX influent and TX effluent (SI Table 3, Figure 3, SI Data). The *msr(E)* gene encodes for macrolide resistance by interfering with the drug’s binding site on bacterial ribosomes (27, 28). Fifteen bacterial hosts were identified for *msr(E)* (SI Table 3). This gene was found on plasmids in *Burkholderiaceae* spp.*, Moraxellaceae* spp., and *Neisseriaceae* spp (Figure 3). In NC influent, taxonomic assignment of bins was able to identify an *msr(E)* host down to the species level: *Acinetobacter johnsonii* in the *Moraxellaceae* family (Figure 3, SI Data). Finally, *tet(C)* encodes for an efflux pump conferring resistance to tetracycline (29–32) and was detected 24 times in the current study with three plasmid-based bacterial hosts identified (SI Table 3).

**Figure 3:**
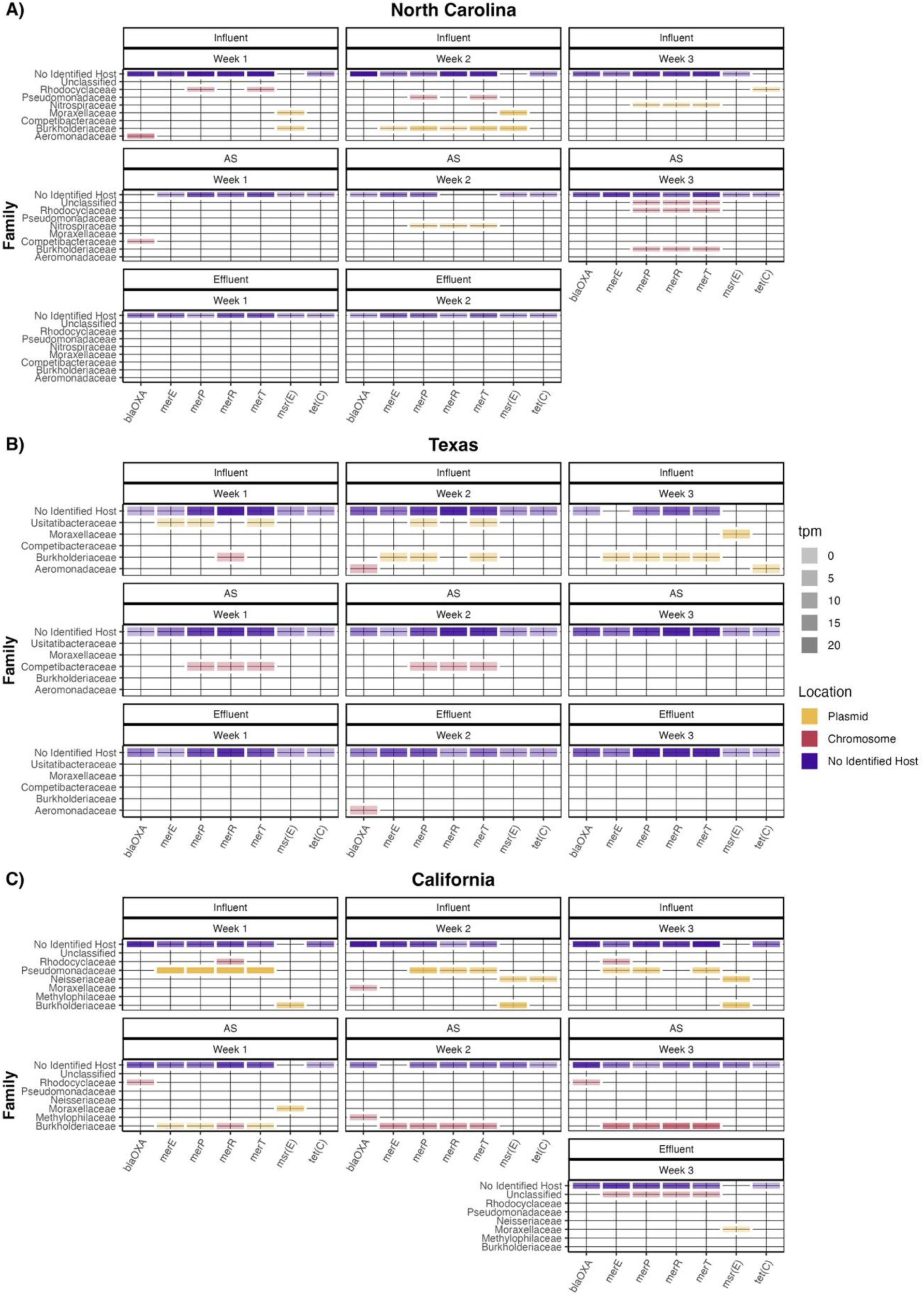
Hosts of the core resistome for each sample collected in A) North Carolina, B) Texas, and C) California. The core resistome is defined as the individual resistance genes detected in every sample collected in this study. Color represents the location in the bacterial genome (either plasmid or chromosome) and the shading represents the transcripts per million (tpm) value. “No Identified Host” were resistance genes annotated on contigs with no informative Hi-C crosslinks or insufficient sequencing depth or quality to identify a bacterial host bin.

### 2.4 Location of Resistance Genes Over Treatment

To understand how the resistome is affected by treatment processes, the host range of ARGs detected in all influent samples of a single WRF (i.e. core influent ARGs) was assessed over treatment (SI Figures 3, 4, and 5, Table 3). There were 20 unique ARGs detected in all NC influent samples (Table 3). Most identified host-ARG relationships in the influent were plasmid based (SI Figure 3, Table 3). Among all shared influent genes in NC, there were only two genes identified in the chromosome: *mph(F)* in *Rhodocyclaceae* and *bla*_OXA_ in *Aeromonadaceae* chromosomes (SI Figure 3). In NC influent, *Burkholderiaceae* spp. contained three different macrolide resistance genes on plasmids: *mph(E)* (33), *msr(E)* (27, 28), and *mph(G)* (34) (SI Figure 3). For ARGs detected in all NC influent samples, microbial hosts were only detected for *bla_OXA_* and *mph(A)* in AS, and these genes were carried in microbial chromosomes (Table 3, SI Figure 3). No microbial hosts were identified in effluent (Table 3, SI Figure 3).

**Table 3:**
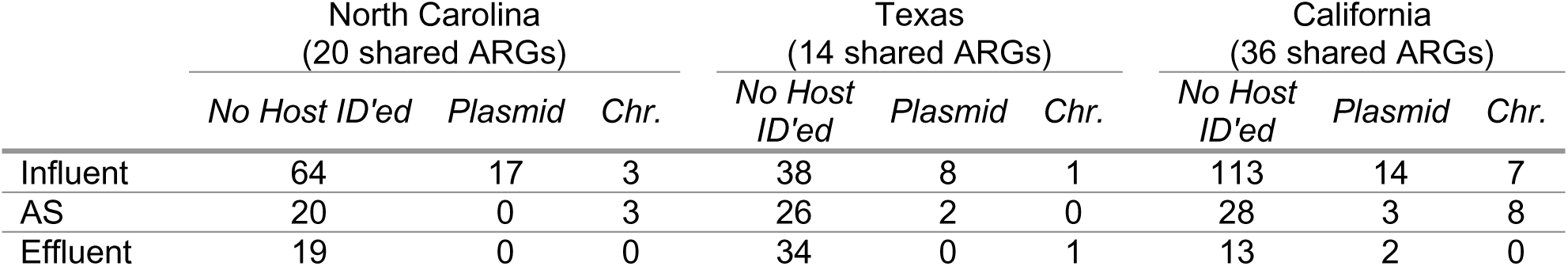
Location in bacterial hosts (plasmid or chromosome (Chr.)) for the core influent ARGs conferring resistance to drugs as they move through the treatment process. Core influent ARGs are resistance genes conferring resistance to pharmaceuticals that were detected in all influent samples within each water reclamation facility. Some ARGs were detected multiple times. Genes with “No Host ID’ed” were annotated in the assembly but had no identified associations with bacterial bins or plasmid after processing Hi-C data.

TX had the fewest ARGs detected in all influent samples (n = 14, Table 3), and most either did not have a host or were on a plasmid (SI Figure 4). The exception to this was *bla*_OXA_, which was detected in the chromosome of an *Aeromonadaceae* spp. in the influent (SI Figure 4). Of the shared ARGs in TX influent samples, most did not have a microbial host identified in TX AS (Table 3). However, two microbial hosts were identified in TX AS, *mph(A)* on a plasmid in a *Polyangiales* spp. and *tet(X2)* on a plasmid in an unclassified *Parvibacillus* (SI Figure 4, SI Data). Only one microbial host was identified in effluent: *bla_OXA_*in the chromosome of an *Aeromonadaceae* spp., the same family it was hosted by in influent (SI Figure 4).

The CA influent had the most unique ARGs detected in all influent samples (n = 36, Table 3). Most identified ARG-bacterial host relationships in influent were on plasmids (Table 3, SI Figure 5). Additionally, numerous bacterial families were detected that hosted ARGs, including *Neisseriaceae* spp., *Enterobacteriaceae* spp., *Burkholderiaceae* spp., and *Bacteroidaceae* spp. (SI Figure 5). *Neisseriaceae* spp. contained plasmids with *msr(E)* macrolide resistance, *sul1* sulfonamide mediated resistance (35), and *tet(C)* mediated tetracycline resistance (31). *Enterobacteriaceae* spp. contained the multidrug resistance genes *acrF* and *emrD* in the chromosome (SI Figure 5) (36, 37). *Burkholderiaceae* spp. contained *msr(E)* and *sul1* on plasmids (SI Figure 5). Finally, *Bacteroidaceae* spp. contained beta-lactamase *cfxA* resistance on a plasmid and *mef(B)* macrolide resistance in its chromosome (38, 39). While most ARGs were identified in bacterial plasmids in influent, in the AS most ARGs were carried on bacterial chromosomes (Table 3, SI Figure 5). In the AS, *sul1* was detected in each sample in the chromosome of *Competibacter phosphatis* (SI Data) but was not detected in this family in the influent. *C. phosphatis* was detected only in the influent in the second week of sampling but without resistance genes. Of the shared ARGs detected in CA influent, only two bacterial hosts were identified in the effluent. *Moraxellaceae* spp. carried a plasmid with *msr(E)* and *Thiobacillaceae* spp. carried a plasmid with *sul1* (SI Figure 5).

Microbial hosts of all resistance genes in effluent were also assessed, with few hosts identified (Table 2, Figure 4). NC effluent had no ARGs with hosts, TX effluent had four ARGs hosted in chromosomes, and CA had 13 ARGs hosted in either plasmids or chromosomes (Table 2, Figure 4). All resistance genes detected in TX effluent conferred resistance to antibiotics, with one of the detected genes also in the core resistome (*bla_OXA_*, Figure 4A). The *bla*_OXA_ gene was found in the chromosome of an *Aeromonadaceae* spp. in TX effluent. *Bacteroidaceae* spp. hosted *lnu(AN2)* mediated lincomycin resistance and *mef(En2)* mediated macrolide resistance (Figure 4A) (40–42). CA effluent had four resistance genes on plasmids and nine in microbial chromosomes (Table 2, Figure 4B). Four of these effluent hosted ARGs conferred drug resistance: *mcr-5, mph(E), msr(E),* and *sul1* (Figure 4B). CA effluent contained *Moraxellaceae* spp. with colistin resistance (*mcr5*) and macrolide resistance (*mph(E)* and *msr(E)*) on plasmids (27, 28, 43, 44). Two ARGs detected in CA influent were detected in hosts from the same sampling week in the effluent: *msr(E)* and *sul1* (SI Figure 5, Figure 4B). The *sul1* gene is in an unclassified *Thiobacillus* species and *msr(E)* is in a *Moraxellaceae* spp. in effluent (Figure 4B).

**Figure 4:**
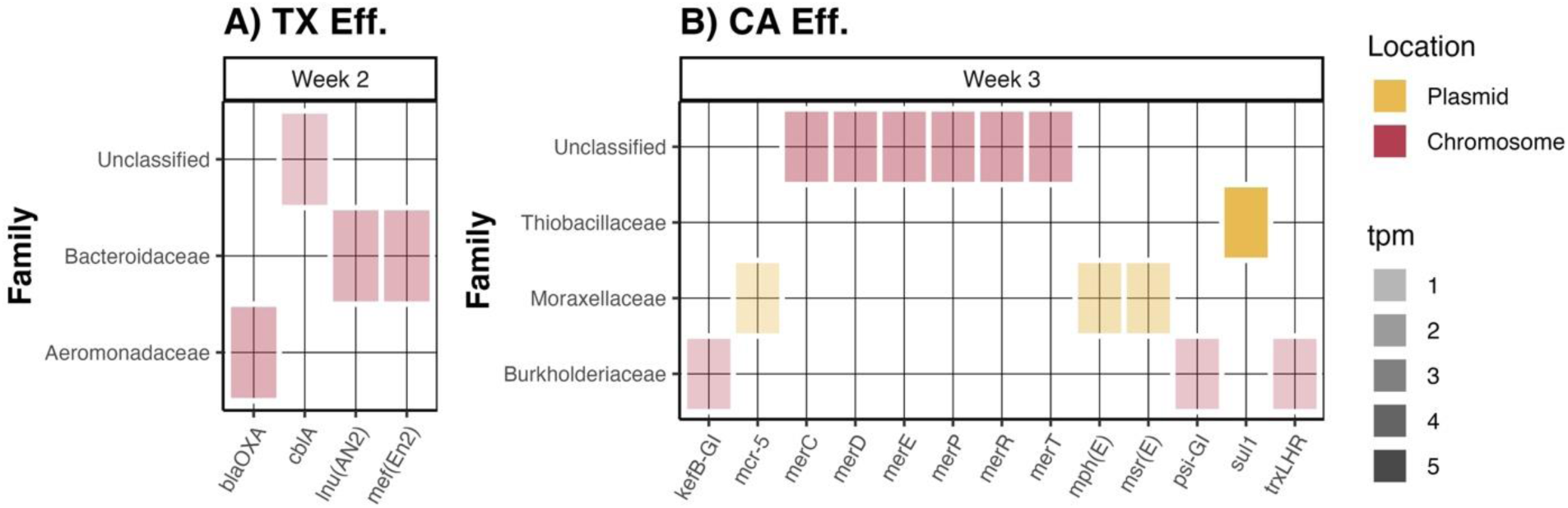
All ARG-host relationships detected by Hi-C contact mapping in effluent samples. Bacterial hosts were identified in A) the Texas sample from week 2 and B) the California sample from week 3. No other effluent samples from other sampling events had bacterial hosts for ARGs. Shading is representative of the transcripts per million (tpm). Unclassified hosts were identified as plasmids and/or bacterial chromosomes crosslinked with resistance genes in the assemblies that did not meet sequencing quality criteria during classification.

### 2.5 Priority resistant families

When considering only ARGs, there were 114 total bacterial hosts identified by Hi-C contact maps (Table 4). More than 50% of the identified relationships involved 4 bacterial families: *Moraxellaceae* spp. (n = 22 ARGs), *Burkholderiaceae* spp. (n = 18 ARGs), *Aeromonadaceae* spp. (n = 13 ARGs), and *Neisseriaceae* spp. (n = 8 ARGs). These families also represented a high relative proportion of the families detected, with average relative abundances ranging from 6.3% to 14.6% (Table 4). While they were not detected in every sample, when present, they represented a relatively high proportion of the total bacterial community (Table 4). Although the bulk of resistance was found in the plasmids in these families from influent samples, there were resistant *Aeromonadaceae* spp. and *Moraxellaeae* spp. detected in the effluent (SI Table 4).

**Table 4:** ARGs conferring resistance to pharmaceuticals (n = 114) hosted by specific bacterial families across all sample types and geographic locations. Mean relative abundance is the average relative abundance of that family across each sample collected where that family was detected at >70% completion and <20% contamination.

| Family | Number of ARGs in family | % total hosted ARGs | Mean Rel. Abun. (95% CI) | Number of samples collected with family detection (% total samples) |
| --- | --- | --- | --- | --- |
| *Moraxellaceae* | 22 | 19.3% | 6.3%<br>(-9.3% - 21.8%) | 8<br>(33%) |
| *Burkholderiaceae* | 18 | 15.8% | 12.8%<br>(-1.8% - 27.4%) | 12<br>(50%) |
| *Aeromonadaceae* | 13 | 11.4% | 9.8%<br>(-0.6% - 20.3%) | 5<br>(21%) |
| *Neisseriaceae* | 8 | 7.0% | 14.6%<br>(-0.7% - 29.9%) | 5<br>(21%) |
| Everything Else | 53 | 46.5% | - | - |

## 3 Discussion

### 3.1 Treatment Stage Affects Location in Microbial Genome but not Overall Resistome

The resistome is highly stable, with all resistance signatures in all samples collected from all three geographic locations dominated by beta-lactam, macrolide, and sulfonamide resistance (SI Table 2, Figure 2A). These classes of antibiotics have been previously identified in environmental and clinical data as being under threat by growing resistance, with high detection of beta-lactam resistance in clinical samples (45) and a high prevalence of macrolide resistance in US wastewater (8). Despite the stability in the resistome, there is little community overlap between influent, AS, and effluent (Figure 1). Influent communities in NC and TX exhibited little similarity week to week, while CA influent was more stable, and AS from all three facilities was highly similar (Figure 1). Stability in the resistome across geography, time, and treatment process despite changes in the underlying communities suggests the resistome is not dependent on the microbial communities and HGT is facilitating resistome stability. Further analysis of the evidence of shared MGEs and plasmids is needed to facilitate full characterization of AMR dynamics in WRFs.

Although the overall resistome was highly stable across the WRFs tested here, resistance gene burden in shotgun data decreased from influent to effluent, with the bulk of reduction occurring from influent to AS (Table 1). However, for sample weeks where the full bioinformatics pipeline was carried out in all three treatment processes, total resistance genes increased from AS to effluent (Table 1). This could indicate the existence of microbial reservoirs in the facility or be an effect of the sampling scheme, which did not take into consideration WRF hydraulic retention times (HRT) and sampling included grab and 24-hour composites. However, previous research does not find that HRT-informed sampling affected pathogen reduction estimates (46); less is known about the impact of sample timing on gene removal. Additionally, other studies have demonstrated the bulk of resistance reduction occurs between influent and AS (12, 47), indicating the patterns observed here are likely not a function of HRT. Further, other published research finds total ARG abundance decreases from influent to effluent while individual ARGs increased from influent to effluent (48), suggesting there are important patterns to be studied at the gene level and in effluent. It is likely that observed patterns were not affected by the use of grab samples in AS and effluent, as AS and effluent are a composite of many time points due to mixing upstream. However, for influent, it has been shown that composite samples better represent pathogen loading in wastewater due to diurnal use of bathroom facilities (49).

The location of resistance genes in microbial genomes is dependent on treatment step, as both total and core resistance genes are more likely to be in a plasmid in influent but in a chromosome in AS (Table 2, SI Table 3). This relative decrease in the proportion of plasmid-based resistance genes from influent to AS has been observed by others (47). It was further expected that the influent resistome would be driven by plasmids, given that influent is representative of the catchment population gut microbiome and the influence of plasmid-based resistance genes in the environment and clinical settings (9, 22, 50–53). While previous work can identify how chromosomal-and plasmid-based resistance genes change through treatment, they are limited by the inability to link plasmid-based genes to their host microbes. The current study was able to successfully link genes on plasmids to hosts due to the Hi-C sample preparation methods.

There were very few microbial hosts identified in effluent samples (Table 2, Figure 4), highlighting both the success of WRF treatment processes at removing resistant bacteria and the difficulties of identifying microbial hosts in matrices with weak genetic signatures. While the facilities used different final disinfection processes (UV in NC, filtration and chlorination in TX, chlorination in CA), it is unclear if effluent host identification is an artifact of sequencing or a function of final disinfection. Existing research does not present a clear answer, as the identified effects of final disinfection on ARB are mixed (54), requiring future research on both ARB and ARGs. However, the identification of ARGs in both microbial chromosomes and plasmids in effluent samples from TX and CA with Hi-C sequencing suggests there are resistant bacteria escaping final disinfection.

### 3.2 Shared Genes Represent High Selective Pressures in WRFs

The resistance genes that were shared among all samples tested were representative of genes experiencing high selective pressure in WRFs. First, four genes in the *mer* operon (*merE, merP, merR,* and *merT*) were identified in all samples that underwent the full bioinformatics pipeline (SI Table 3). Although mercury and other heavy metals were not measured here, heavy metals in WRFs have been found to be at high enough concentrations to increase plasmid conjugation frequency and drive resistance for various resistance genes (13, 55–58). There were also three commonly identified ARGs found in all samples analyzed (SI Table 3): *bla_OXA_*, which is one of the most commonly identified carbapenem resistance genes across sample type (59–61), and the macrolide resistance gene *msrE* and the tetracycline resistance gene *tetC* which were both detected by P. Munk et al. (8) in 757 wastewater samples collected from 101 countries. Antibiotics were not analyzed here, but antibiotics are commonly detected in WRFs at levels that drive resistance (10). Future work should aim to quantify heavy metal and antibiotic concentrations to better understand drivers of AMR dynamics with Hi-C sequencing.

### 3.3 Critical Microbial Families and Species Identified as ARG Hosts

Although species level annotation of microbial resistance gene hosts overall was limited, there were a few notable species-level detections. *A. caviae* with chromosomal *bla_OXA_* beta-lactam resistance was detected in NC and TX influent and TX effluent (Figure 3, SI Data). Beta-lactam resistant *A. caviae* was previously isolated from advanced water purification facilities (62). Detection in influent and effluent from two of the WRFs sampled here suggests this resistant bacterium is common in the population and can survive the wastewater treatment processes presented here and advanced water treatment processes previously reported. A chromosomal *msr(E)* encoded macrolide resistant *A. johnsonii* bacterium was additionally detected in NC influent (Figure 3, SI Data). *A. johnsonii* is primarily a commensal bacterium but can cause opportunistic catheter-related bloodstream infections (63, 64). Although *A. johnsonii* has been proposed as a reservoir of ARGs (63), there is limited evidence it carries extensive macrolide resistance. This detection by Hi-C could represent either a new resistotype or further support the hypothesis that it is an ARG reservoir. Bacterial species involved in the nitrogen cycle, *Nistrospira defluvii* (*Nitrospiraceae* family) and *Comamonas denitrificans* (*Burkholderiaceae* family), were identified with plasmid-based *mer* mercury resistance and *msr(E)* macrolide resistance in NC influent and AS (Figure 3, SI Data), indicating resistome dynamics are not limited to clinical bacteria in these facilities.

There were additionally several microbial-ARG host relationships identified in AS and effluent that were not detected in influent. Each AS sample collected in CA contained *C. phosphatis* with *sul1* chromosomal sulfonamide resistance (SI Data, SI Figure 5), despite a lack of this relationship in influent. While there are likely other sources of *sul1* in the WRF, recurrent detection suggests persistence of this ARG-microbial relationship independent of influent bacteria. Additionally, among the resistance genes detected in TX effluent, only *bla_OXA_* was in the same microbial family in effluent as influent (Figure 3B, Figure 4A). Further in CA, the only two ARGs identified with microbial hosts in effluent that were also detected in influent (*sul1* and *msr(E)*) were in plasmids in different microbial hosts in effluent than in influent. This could suggest either sporadic detection of resistant bacteria throughout the WRF with Hi-C or be representative of AMR dynamics in the facilities. Even though species level identification was limited, as shown here, Hi-C can be used to better understand the resistome without isolating and culturing resistant bacteria.

A majority of ARGs for pharmaceuticals were hosted by four bacterial families: *Moraxellaceae* spp., *Burkholderiaceae* spp., *Aeromonadaceae* spp., and *Neisseriaceae* spp. (Table 4). Further, a majority of ARGs with bacterial hosts were detected in plasmids (SI Table 4), suggesting the risk for HGT is high. The Hi-C contact maps indicate that these four bacterial families are AMR reservoirs. Both *Moraxellaceae* spp. and *Aeromonadaceae* spp. were previously identified as AMR reservoirs in wastewater treatment systems (21, 65). Furthermore, the presence of resistant and intact *Aeromonadaceae* spp. and *Moraxellaceae* spp. in WRF effluents (Figure 4, SI Table 4) emphasizes the need to understand drivers of these bacterial families to reduce environmental AMR exposure risks and reduce HGT of the genes they carry.

### 3.4 Conclusions

The current study identified numerous novel ARG-microbial relationships and potential reservoirs of AMR in water reclamation facilities in three geographically distinct areas. *Moraxellaceae* spp., *Burkholderiaceae* spp., *Aeromonadaceae* spp., and *Neisseriaceae* spp. were together responsible for more than 50% of microbial-ARG host relationships. Commensal, pathogenic, and environmental bacterial species were detected as ARG hosts across wastewater treatment, suggesting Hi-C elucidates ARGs across bacterial families. Further, the location of ARGs in the microbial genome was dependent on the treatment step, as ARGs were more likely to be in microbial plasmids in influent and in microbial chromosomes in AS. While total ARGs decreased from influent to effluent, there were increases in ARGs from AS to effluent in individual sample weeks and resistant bacteria were detected in effluent. Overall, more research into the specific drivers of ARG transfer in WRFs is needed to inform risk assessments.

Hi-C contact mapping is a powerful shotgun sequencing tool to identify ARG hosts in complex environmental matrices. Hi-C and other proximity ligation sequencing methods are the only nonspecific methods to definitively link MGEs with bacterial hosts by physically crosslinking plasmids with bacterial chromosomes before cell lysis. Hi-C has the same limitations as shotgun sequencing in that metagenomic annotation is dependent on the databases utilized and sequencing quality and depth affect final results. Therefore, there are likely unidentified relationships in these samples that would benefit from more targeted single cell sequencing techniques like epicPCR and long-read sequencing with unique molecular identifiers (20, 66). Although the sample size in the current study is relatively small, given the novelty of proximity ligation techniques, the results presented advance understanding of the resistome in WRFs, particularly for resistance genes carried on plasmids.

## Data Availability

All data produced in the present study are available upon reasonable request to the authors

## 4. Acknowledgments

This work would not have been possible without the support of our collection teams at the WRFs in NC, TX, and CA and Hira Waheed who helped collect and prepare samples in NC. We also thank the PhaseGenomics team for their support interpreting the Hi-C contact maps and running the deconvolution platform. The authors acknowledge the Center for Advanced Research Computing (CARC) at the University of Southern California for providing computing resources that have contributed to the research results reported within this publication (URL: https://carc.usc.edu).

## 6 Materials and Methods

### 6.1 Sample Collection and Concentration

Samples were collected weekly during the same weeks from three WRFs in CA, TX, and NC. All three WRFs discharge the effluent into local surface waters. Primary influent was either grab-sampled in the morning (NC) or collected as a 24-hour composite sample (CA and TX). Activated sludge (AS) was grab-sampled from all three WRFs. Effluent was collected after disinfection with either UV (NC) or chlorine (CA and TX) via grab sampling (NC and TX) or a 24-hour composite (CA). Sample collection volumes and dates are in SI Table 5. Samples collected in TX and NC were shipped overnight with refrigeration to the ReWater Center at the University of Southern California. All samples were then stored at 4°C until processing.

Primary influent (50mL) and AS (2.0mL) from all three WRFs were centrifuged at 12,000 x *g* and 4°C for 10 minutes (21). The supernatant was then removed and pellets stored at 4°C. Effluent (500mL) was centrifuged at 3500 x *g* and 4°C for 30 minutes in 50mL conical centrifuge tubes. The supernatant was removed, and pellets were combined. The combined pellets were then recentrifuged and the supernatant removed for a single pellet from 500mL equivalent effluent volume. This pellet was stored at 4°C. All processing for all three sample types was carried out in duplicate, with one replicate used for Hi-C crosslinking and the other for DNA extraction and shotgun sequencing.

### 6.2 Hi-C Preparation

Hi-C crosslinking was carried out within one week of collection. One sample replicate from each location, sample type, and sampling date was processed using the ProxiMeta^TM^ Hi-C Kit following protocol v4.5.4 (Phase Genomics, Seattle, WA, USA) with the following alterations to crosslinking to account for solids content. Samples were crosslinked with 10mL of a chilled 1X PBS with 1% formaldehyde solution for 10 minutes at room temperature. Crosslinking was quenched with 527uL of 2.5M glycine for five minutes. Samples were then transferred to ice for 10 minutes with periodic agitation. After chilling, samples were centrifuged at 3500 x *g* and 4°C for 10 minutes. The supernatant was discarded, and pellets resuspended in 10mL of chilled 1X PBS. Resuspended samples were centrifuged again at 3500 x *g* and 4°C for 10 minutes. The supernatant was discarded, and pellets resuspended in 1.0mL of chilled 1X PBS. Samples were centrifuged at 800 x *g* and 4°C for 5 minutes, supernatant discarded, and crosslinked pellets stored at -80°C until further processing (67). After crosslinking, processing followed the protocol as written.

### 6.3 DNA Extraction

DNA was automatically extracted from remaining pellet replicates using the Maxwell ^®^ Blood DNA Kit using the Maxwell ^®^ RSC 48 instrument (Promega Corp., Madison, WI, USA). Cell lysis was modified to account for additional solids in wastewater samples. Briefly, 300uL of the included lysis buffer and 0.5g of sterilized zirconia beads (BioSpec Products Inc, Bartlesville, OK) were added to the pellets. Samples underwent bead beating at 2400 RPM for two minutes using a Mini-BeadBeater 96 (BioSpec Products Inc, Bartlesville, OK) followed by centrifugation at 15,000 x *g* for 5 minutes. 200uL of supernatant was transferred to a sterile 1.5mL microcentrifuge tube. 200uL of lysis buffer was added to the bead tube. The whole process was repeated twice for three total rounds resulting in 600uL total of cell lysate. Proteinase K was then added to the cell lysate (60uL) and mixed by vortexing. Cell lysate was incubated at 56°C for 30 minutes. The extraction protocol then proceeded as written with final elution in 40uL of nuclease-free water.

### 6.4 Sequencing and Bioinformatics Analysis

DNA extracts were sent to the Oklahoma Medical Research Foundation Clinical Genomics Core (Oklahoma City, OK) for library preparation and shotgun sequencing. Shotgun libraries were prepared using a Watchmaker DNA Library Prep Kit (Watchmaker Genomics Boulder, CO). Indexed Hi-C sample libraries were sent to the Duke Sequencing and Genomic Technologies Core Facility (Durham, NC) for sample pooling and shotgun sequencing. Both the shotgun and Hi-C libraries were sequenced at a depth of 100 million reads/sample using one lane of an Illumina NovaSeq X Plus (Illumina San Diego, CA).

Shotgun sequencing libraries were assembled utilizing the USC Center for Advanced Research Computing’s high-performance computing systems. Initial quality control analysis of the fastq files was performed using FastQC (68). Low-quality reads and adapters were filtered and trimmed using bbDuk (69). Contigs were assembled using metaSPADES (70). Mean contig length for all samples that underwent Hi-C deconvolution was obtained using the quast.py command from QUAST (v5.0.3) and are in SI Table 1 (71, 72). The commands with all flags are in Supplemental Appendix B.

Hi-C deconvolution, binning, and annotation was performed at Phase Genomics (Seattle, Washington, USA) using the ProxiMeta platform following similar methods as described in M. O. Press et al. (73) and A. Risely et al. (22). In brief, Hi-C reads were aligned to the metagenomic assemblies with the –5SP flag using BWA-MEM (74), the alignments were filtered using the –F 2304 flag with SAMtools (75), and assemblies were binned using the ProxiMeta platform (73, 76). In contrast with binning traditional shotgun sequencing data, Hi-C binning incorporates the crosslinked DNA to bin the assemblies into genomes, plasmids, and viruses rather than relying solely on sequence similarity (73). Average coverage of the Hi-C generated bins to the trimmed shotgun sequencing reads was calculated using the CoverM genome command with the -p flag using BWA-MEM and -m flag using tpm (77).

The Genome Taxonomy Database Toolkit (GTDB-tk, v2.4.0) and database v220 were used to classify bin taxonomy (78). Plasmid contigs were identified by blastn of all contigs using the PLSD database (v2020_06_29) and the Hi-C data were used to bin plasmids into pMAGs (79, 80). AMR genes were annotated from the metagenomic assemblies using AMRFinderPlus (v3.10.5). The gene annotations were cross-referenced with all MAGs and host-associations and were assigned to their origin by using a binary matrix. MAGs were generated by filtering assembled bins for > 70% completion and <20% contamination using the dplyr R package (81). Three effluent samples (one from NC and two from CA) were not able to undergo the complete Hi-C pipeline because there was not sufficient mapping of the Hi-C reads to the assemblies, and these samples were removed from subsequent analysis. Raw sequences are deposited in the National Center for Biotechnology Information’s Sequence Read Archive database at project number PRJNA1295499.

### 6.5 Data Management and Statistical Analysis

All statistical analysis and data manipulation was carried out using R (82), RStudio (83), and affiliated packages. Data were imported into RStudio using readr (84). Data were manipulated using the dplyr and tidyverse packages (81, 85). Plots were generated using ggplot2 (86), ggh4x (87), scico (88), pals (89), ggpubr (90), cowplot (91), and ggbiplot (92). Relative abundance of the MAGs, antibiotic resistance gene, and total resistance to antibiotic class was calculated by adjusting the transcripts per million (TPM) reads to the relative fraction for each target (Equation 1).

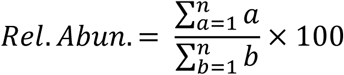

*Equation 1: Relative abundance calculations where a = tpm for a single MAG, antibiotic resistance gene, or antibiotic class and b = tpm for all MAGs or antibiotic resistance genes*

Although transcripts were not analyzed in this study, TPM is a metric that normalizes shotgun sequencing data to the read length and sequencing depth to allow for comparability between samples and is comparable to relative abundance. Principal component analysis (PCA) with centering and scaling following Hellinger transformation of the data was carried out using the prcomp function in the R Stats Package, the labdsv package (93), and the factoextra package (94).

